# Temporal Dynamics of the Adult Female Lower Urinary Tract Microbiota

**DOI:** 10.1101/2020.03.06.20032193

**Authors:** Travis K. Price, Birte Wolff, Thomas Halverson, Roberto Limeira, Linda Brubaker, Qunfeng Dong, Elizabeth R. Mueller, Alan J. Wolfe

## Abstract

Temporal dynamics of certain human microbiotas have been described in longitudinal studies; variability often relates to modifiable factors or behaviors. Early studies of the urinary microbiota preferentially used samples obtained by transurethral catheterization to minimize vulvo-vaginal microbial contributions. Whereas voided specimens are preferred for longitudinal studies, the few studies that reported longitudinal data were limited to women with lower urinary tract (LUT) symptoms, due to ease of accessing a clinical population for sampling and the impracticality and risk of collecting repeated catheterized urine specimens in a non-clinical population. Here, we studied the microbiota of the LUT of non-symptomatic, pre-menopausal women using mid-stream voided urine (MSU) specimens to investigate relationships between microbial dynamics and personal factors. Using 16S rRNA gene sequencing and a metaculturomics method called Expanded Quantitative Urine Culture (EQUC), we characterized the microbiotas of MSU and peri-urethral swab specimens collected daily for approximately three months from a small cohort of adult women. Participants were screened for eligibility, including ability to self-collect paired urogenital specimens prior to enrollment. In this population, we found that measures of microbial dynamics related to specific participant-reported factors, particularly menstruation and vaginal intercourse. Further investigation of the trends revealed differences in composition and diversity of LUT microbiotas within and across participants. These data, in combination with previous studies showing relationships between the LUT microbiota and LUT symptoms, suggest that personal factors relating to the genitourinary system may be an important consideration in the etiology, prevention, and/or treatment of LUT disorders.

**IMPORTANCE:** Following the discovery of the collective human urinary microbiota, important knowledge gaps remain, including the stability and variability of this microbial niche over time. Initial urinary studies preferentially utilized samples obtained by transurethral catheterization to minimize contribution from vulvo-vaginal microbes. However, catheterization has the potential to alter the urinary microbiota; therefore, voided specimens are preferred for longitudinal studies. In this report, we describe the microbial findings obtained by daily assessment over three months in a small cohort of adult women. Similar to vaginal microbiotas, we found that lower urinary tract (LUT) microbiotas are dynamic with changes relating to several factors, particularly menstruation and vaginal intercourse. Our study shows that LUT microbiotas are both dynamic and resilient. It also offers novel opportunities to target LUT microbiotas for preventative or therapeutic means, through risk and/or protective factor modification.

## INTRODUCTION

The microorganisms of our bodies are collectively known as the human microbiota^1^. These communities of bacteria can influence many aspects of health and disease. The balance of benefit versus harm depends largely on the overall state of the microbiota in terms of their distribution, diversity, and composition^2^. Understanding the variables associated with microbial changes is an important first step to intentional modulation of the microbiota. Compared to higher biomass microbial niches, such as the gut, little is known about the temporal dynamics of the female urinary microbiota.

Temporal changes in the vaginal microbiota have been described. Overall, the vaginal microbiota shows low microbial constancy and high species turnover over time, but the dynamics vary widely among individuals^3 4^. Vaginal microbiotas can also be resilient^5^, meaning they return to a baseline state following disruption. Alterations in vaginal microbiotas relate to vaginal health [e.g., bacterial vaginosis (BV)]^5 6^ and personal factors (e.g., menstruation, sexual intercourse, contraception, pregnancy, menopausal status)^3 5 7 8 9 10 11 12 13 14^. Incidence of BV can also fluctuate^15^ and relates to personal factors^16 17 18 19 20 21^. Altogether, these data suggest possible interplay amongst personal factors and practices, microbial dysbioses and vaginal health.

Likewise, urinary tract infection (UTI) risk in women is associated with similar personal factors^22 23 24 25 26^. These data, in combination with the discovery of resident microbiotas in the bladders of women^27 28 29 30 31 32^, provide evidence to propose similar interplay for the female lower urinary tract (LUT), consisting of the bladder and urethra, which are in close proximity to the vagina. To date, only one study has assessed the urinary microbiota longitudinally. In males, Nelson *et al*. showed that voided urine specimens collected at one-month intervals were significantly more similar within a participant than between participants^33^, suggesting microbial stability. They found that some bacterial taxa (e.g., *Propionibacterium* and *Lactobacillus*) had long durations of colonization, while others (e.g., *Corynebacterium, Anaerococcus, Staphylococcus*, and *Prevotella*) were more infrequent^33^. Furthermore, BV-associated taxa (e.g. *Mycoplasma, Ureaplasma*, and *Sneathia*) were detected only in sexually experienced males^33^. These data show that urinary microbiotas in males are dynamic and may relate to personal factors. Whether this is true for the female LUT microbiota, and whether daily assessment of the microbiota will provide further clarity, remain unclear and serve as the primary objective of this study.

In this study, we characterized the microbiotas of mid-stream voided urine (MSU) and peri-urethral swab in specimens collected daily for three months from pre-menopausal adult women without LUTS. This is a secondary analysis of a previously described study^34^, where we measured the change in the ratio of urinary pathogens and *Lactobacillus spp*. within the LUT in response to oral probiotic use. In this double-blinded randomized controlled trial, we found no effect of oral probiotic use on the LUT microbiota^34^. Here, our aims were to describe the longitudinal microbiotas of MSU and peri-urethral swab specimens in this participant population, and to determine whether correlations existed between temporal changes in the microbiotas and participant-reported biological and behavioral factors.

We found that, similar to vaginal microbiota, LUT microbiota are dynamic and are associated with specific biological and behavioral factors, particularly menstruation and vaginal intercourse. Our study provides the first descriptive analysis of temporal changes in the LUT microbiota. It shows that these microbiota are relatively stable during health. They can be modulated temporarily by certain behavioral and biological factors. But, the LUT microbiota of young healthy women are remarkably resilient. It also offers novel opportunities to target the LUT microbiota for preventative or therapeutic means, through lifestyle modifications.

## RESULTS

### Study Design and Patient Demographics

Because post-urethral (e.g. vulva and/or vaginal) microbes are often present in MSU samples^35 36^ it was essential to include a control for specimen quality. Thus, we used a peri-urethral swab to measure specimen collection compliance. We prescreened participants, seeking those whose peri-urethral swab microbiota and MSU microbiota differed substantially, allowing us to make conclusions that exclusively relate to the LUT microbiota rather than to those of the peri-urethra.

We screened 12 participants for eligibility by calculating Bray-Curtis Dissimilarity Index values between microbiotas of paired MSU and peri-urethral specimens collected over three consecutive days. The Bray-Curtis Dissimilarity Index is used to quantify the compositional dissimilarity between two sites. This index produces values between 0 and 1, with a value of 0 assigned to completely similar specimens. Four participants did not meet eligibility criteria (**Supplemental Table 1**).

The other 8 participants entered into the study, which had 3 phases (**Figure 1**). During Phase I (days 1 – 20) and Phase II (days 21-60), all participants collected daily MSU and peri-urethral specimens and completed a questionnaire. During Phase II, participants were randomized to take an oral probiotic or placebo. This part of the study has been published^23^. During Phase III (days 61 – 95), all participants collected MSU and peri-urethral specimens and completed the questionnaire on a daily basis from days 61 – 74, followed by weekly collection and questionnaire completion from days 74 – 95. Specimens were delivered each morning for laboratory analysis.

**Figure 1.**
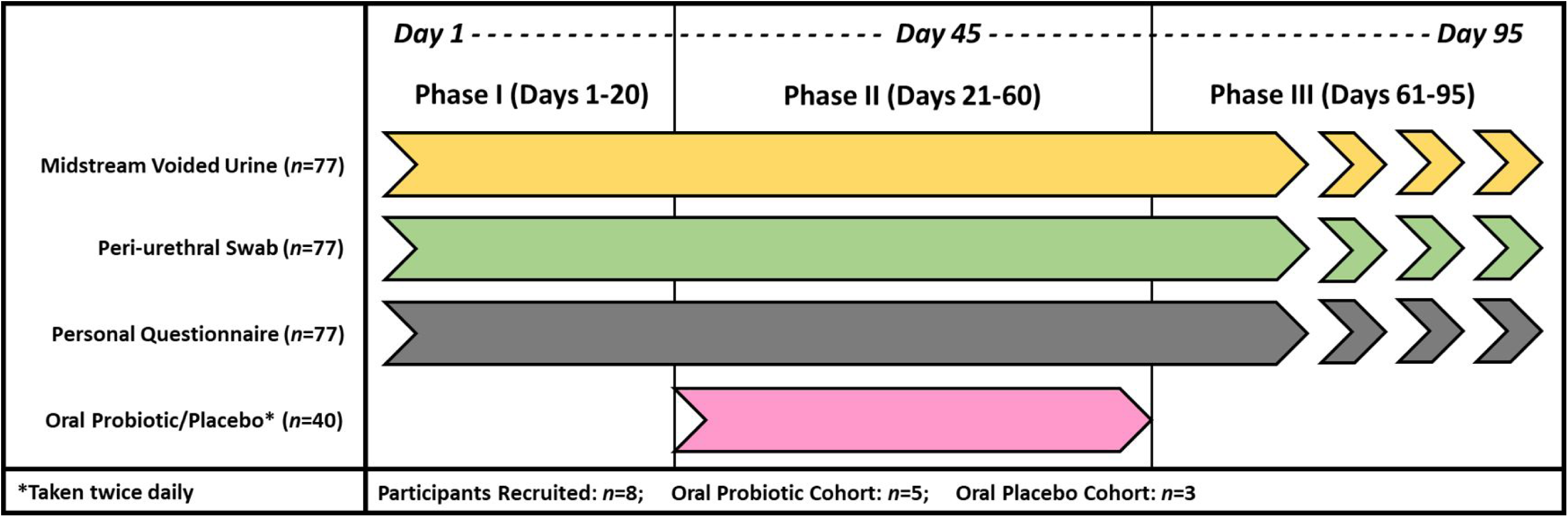
ProFUM Clinical Study Design Outline. Midstream Voided Urine (MSU) and peri-urethral swab specimens were self-collected daily from 8 participants from Days 1 – 74, followed by weekly collection through Day 95. Daily questionnaires were completed on each of these days. The study design was divided into three phases: Phase I (Days 1 – 20), Phase II (Days 21 – 60), and Phase III (Days 61-95). During Phase II, the participants were randomized (2:1) to take an oral probiotic or placebo twice daily. This assignment was double-blinded.

None of the participants had symptoms of urinary incontinence or a prior history of gynecological surgery, kidney stones, or recurrent urinary tract infections (**Table 1**).

**Table 1.** Demographics and Clinical Characteristics of the Participants.

| **ProFUM Participant Demographics** | **Total cohort (n=8)** |
| --- | --- |
| **Age (years), mean (SD)** | 29 (± 5) |
| **Race/Ethnicity** |  |
| Caucasian/White | 5 (62.5%) |
| Asian | 2 (25%) |
| Black/African American | 1 (12.5%) |
| **BMI, mean (SD)** | 24.6 (± 6.2) |
| **Prior pregnancy** | 1 (12.5%) |
| Vaginal delivery | 1 (12.5%) |
| **Menstrual cycle** |  |
| Regular (every 20-40 days) | 5 (62.5%) |
| Irregular | 3 (37.5%) |
| None | 0 (0%) |
| **Menstrual hygiene product use:** |  |
| -Tampons | 5 (62.5%) |
| -Pads | 4 (50%) |
| -Menstrual cups | 2 (25%) |
| -None | 0 (0%) |
| **Sexually active** | 8 (100%) |
| **Frequency of sexual activity** |  |
| Daily | 1 (12.5%) |
| Weekly | 2 (25%) |
| Monthly | 3 (37.5%) |
| Not in past year | 2 (25%) |
| **Type of sexual activity (in past year):** |  |
| **(N=6)** |  |
| -Vaginal (penetrative with penis) | 6 (100%) |
| -Vaginal (penetrative with toy/fingers) | 6 (100%) |
| -External stimulation | 2 (33.3%) |
| -Cunnilingus (receiving) | 6 (100%) |
| -Anal (penetrative) | 0 (0%) |
| **Current sexual partners** |  |
| One male partner | 6 (75%) |
| Multiple male partners | 1 (12.5%) |
| No current partners | 1 (12.5%) |
| **Method of birth control:** |  |
| -Condom use | 2 (25%) |
| -Intrauterine device (IUD) | 4 (50%) |
| -Oral contraceptive | 2 (25%) |
| **Hygiene** |  |
| Showers (weekly), mean (SD) | 6 (± 1) |
| Baths (weekly), mean (SD) | 1 (± 1) |
| **Bowel movements (weekly), mean (SD)** | 8 (± 3) |
| **Diet** |  |
| Follows a special diet | 1 (12.5%) |
| Alcohol consumption | 8 (100%) |

### Microbiota Characteristics

To evaluate if the MSU and peri-urethra microbiotas remained distinct in the participants over the course of the study, we compared their compositions using the Bray-Curtis Dissimilarity Index. The microbiotas of the paired (i.e., collected on same day) MSU and peri-urethral swab specimens remained distinct (**Supplemental Table 2**). Similarly, principal coordinate analyses (PCoA) showed that, in general, the microbiotas of the MSU and peri-urethra in the individual participants remained distinct over the course of the 90 days study (**Figure 2)**.

**Figure 2.**
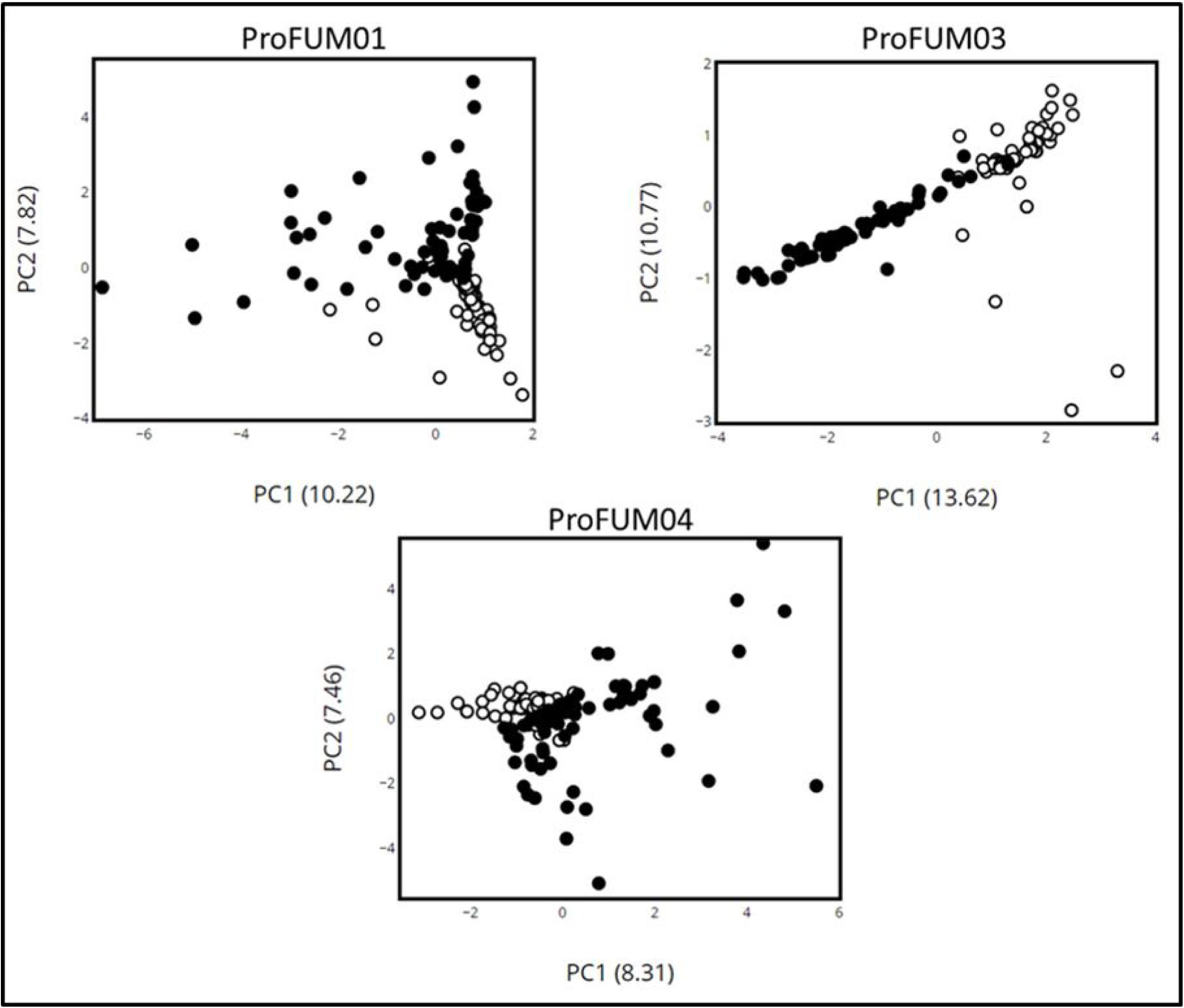
Principal Coordinate Analyses of Microbiota of Specimens from Participants ProFUM01, ProFUM03, and ProFUM04. Analysis was done using MSU (closed circles) and peri-urethral (open circles) microbiota data collected from three participants: ProFUM01 (*top left*), ProFUM03 (*top right*), and ProFUM04 (*bottom*). Graphs plot the 1^st^ and 2^nd^ principal coordinates of the data. Percent of total variance explained by each principal coordinate is shown in the parentheses.

### Qualitative and Quantitative Description of the Longitudinal LUT Microbiota and Microbiome

We evaluated the composition of the participants’ LUT microbiotas using a modified version of the Expanded Quantitative Urine Culture (EQUC) protocol. In parallel, the microbiotas of two participants were evaluated using 16S sequencing (**Figure 3**). In particular, the data for participant ProFUM7 show that EQUC and 16S sequencing provide similar results.

**Figure 3.**
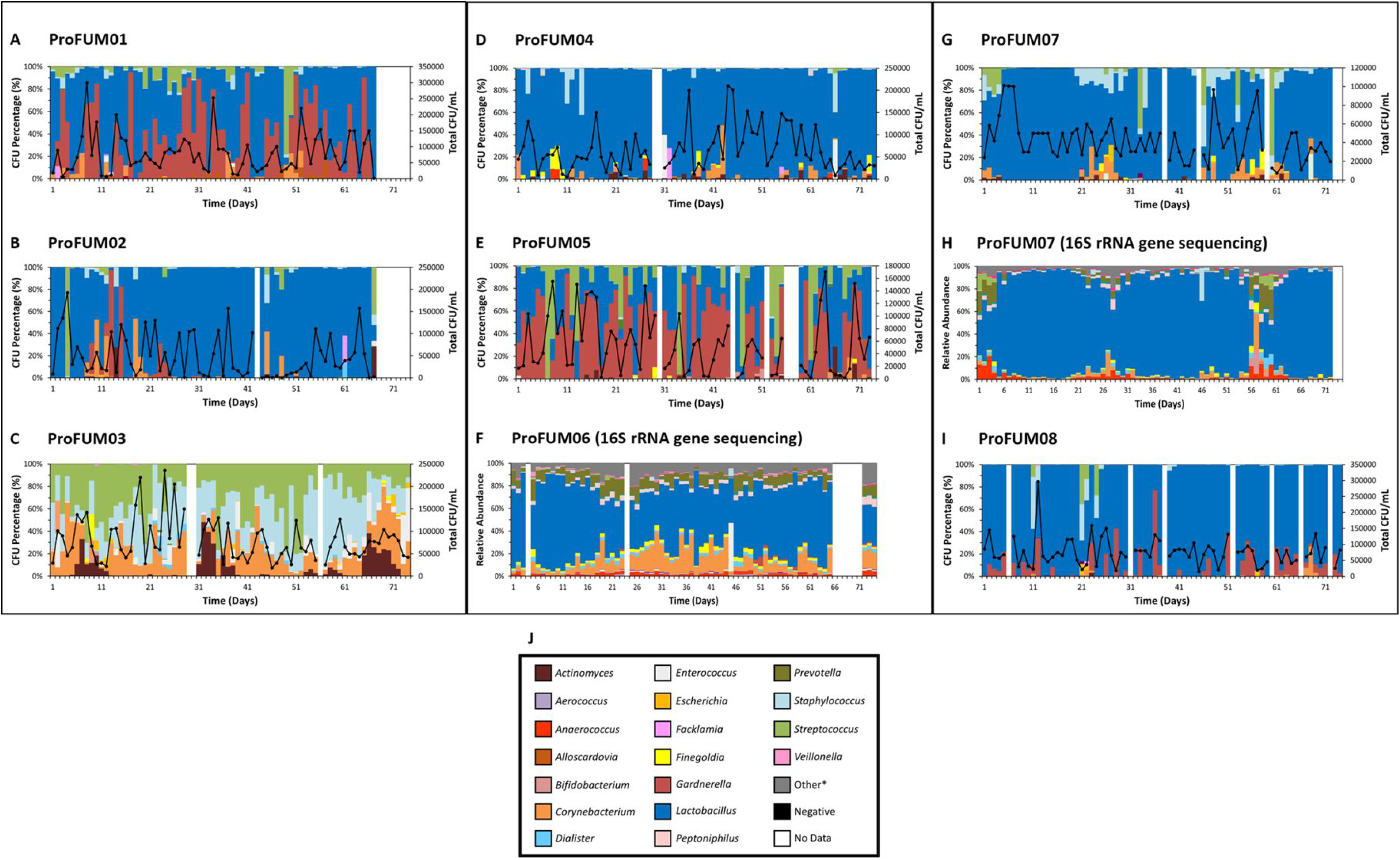
Microbiota Profiles of MSU Specimens from all ProFUM Participants. Microbiota profiles are shown as stacked bar graphs depicting the relative abundance (y-axes) of various genera over time in chronological order (x-axes) from MSU specimens. Bars that appear ‘white’ refer to days where no specimen was collected, received, or stored. A legend containing the most common genera is found in panel **J**. ‘Other’ refers to the combined relative abundance for all taxa not included in the 20 most abundance taxa. Data were generated using modified EQUC (panels **A, B, C, D, E, G**, and **I**) or 16S rRNA gene sequencing (panels **F** and **H**). *Other: only used for panels **F** and **H**

Three general patterns were observed: (i) *Lactobacillus*-predominance (ProFUM02, ProFUM04, ProFUM07, ProFUM08) (ii) alternating *Lactobacillus-* and *Gardnerella*-predominance (ProFUM01, ProFUM05) and (iii) changing ratios of *Streptococcus, Staphylococcus* and *Corynebacterium* (ProFUM03). *Lactobacillus*-predominance could be restricted to a single (ProFUM07) or alternating (ProFUM 02, ProFUM04, ProFUM08) species. Less prevalent taxa were observed in each participant; some were associated with personal factors (see below). CFU/mL varied across five orders of magnitude with a maximum of approximately 250,000. A detailed description of each participant and their MSU composition is presented (**Supplemental Table 3**).

To quantify the stability for both the microbiotas and microbiomes of the MSU and peri-urethral specimens, we used the Jensen-Shannon Divergence (JSD) statistic, a method of measuring the similarity between two probability distributions. Median JSD values were lower for MSU specimens compared to peri-urethral (**Supplemental Figure 1A)**, showing that the peri-urethral microbiota is more variable within an individual than is the MSU microbiota. However, the range of JSD values for the MSU microbiotas was large, indicating that periods of heightened variability exist (**Supplemental Figure 1B**).

### Menstruation and LUT Microbiota Variability

Menstruation was reported by all eight participants. Three participants reported having irregular menstrual cycles, while five reported having regular cycles defined as once every 20-40 days (**Table 1**). Five participants reported using a form of contraception/birth control other than condom use (**Table 1**). JSD values were higher for the MSU microbiotas during menstruation for most participants (**Table 2**). In contrast, JSD values for the peri-urethral microbiotas were not significantly higher during menstruation, except for participant ProFUM02 (*p*<0.001) (**Table 2**). These data show that a significant association exists between menstruation and variable MSU microbiotas, but not between menstruation and peri-urethral microbiotas.

**Table 2.** Association between Jensen-Shannon Divergence Values for MSU and Peri-urethral Microbiota and Participant-reported Menstruation.

| Menstruation |  |  |  |  |  |  |
| --- | --- | --- | --- | --- | --- | --- |
| Participants | MSU Microbiota<br>(Median JSD Values) |  |  | Peri-urethral Microbiota<br>(Median JSD Values) |  |  |
|  | "Yes" (n) | "No" (n) | p-value | "Yes" (n) | "No" (n) | p-value |
| **ProFUM01** | 0.200 (15) | 0.145 (52) | 0.109 | 0.355 (15) | 0.324 (50) | 0.729 |
| **ProFUM02** | 0.316 (7) | 0.127 (59) | 0.039 | 0.377 (7) | 0.172 (55) | <0.001 |
| **ProFUM03** | 0.137 (18) | 0.100 (53) | 0.041 | 0.127 (16) | 0.112 (47) | 0.313 |
| **ProFUM04** | 0.122 (12) | 0.059 (60) | 0.023 | 0.320 (10) | 0.253 (56) | 0.088 |
| **ProFUM05** | 0.101 (3) | 0.129 (64) | 0.989 | 0.240 (3) | 0.252 (57) | 0.873 |
| **ProFUM06 (16S)** | 0.094 (7) | 0.072 (59) | 0.540 | - | - | - |
| **ProFUM07** | 0.078 (41) | 0.052 (28) | 0.014 | 0.246 (41) | 0.322 (28) | 0.095 |
| **ProFUM07 (16S)** | 0.070 (43) | 0.048 (29) | <0.001 | - | - | - |
| **ProFUM08** | 0.098 (19) | 0.068 (48) | 0.047 | 0.227 (19) | 0.297 (47) | 0.051 |

Median JSD values shown for MSU (*left*) and peri-urethral (*right*) microbiota on days when the participant reported ("Yes") or did not report ("No") menstruation. Mann-Whitney U test used to determine significance. *p*-value < 0.05 is significant (green).

To determine the nature of MSU microbiota variability associated with menstruation, we assessed for differences in microbial composition and diversity. Median alpha-diversity measures during menstruation were significantly higher for most participants (**Supplemental Table 4**), particularly those participants who previously showed high MSU microbiota variability during menstruation (**Table 2**). We also observed numerous differences in the frequency of detected taxa during menstruation. **Supplemental Table 5A** lists the bacterial taxa that had significantly different frequencies of detection between menstruation and non-menstruation days. Five participants (ProFUM01, ProFUM02, ProFUM03, ProFUM04 and ProFUM07) had at least one bacterial taxon that was statistically significant; three participants (ProFUM08, ProFUM05, and ProFUM06) did not; the latter two previously showed no difference in alpha-diversity (**Supplemental Table 4**) or JSD values (**Table 2**) during menstruation. Overall, changes in MSU microbial stability and composition were associated with menstruation, but participants showed individualized trends.

### Sexual Activity and LUT Microbiota Variability

Sexual activity was reported by six participants (**Supplemental Table 6**). Of these six participants, penetrative vaginal intercourse was reported by all at least once, and was the most commonly reported form of sexual activity. For the applicable participants (ProFUM06, ProFUM07, and ProFUM08), condom use was reported for 100% of the reported instances of vaginal intercourse. One participant, ProFUM07, reported being with the same male partner for all instances of vaginal intercourse (5/5, 100%), while ProFUM08 reported different male partners for each instance of vaginal intercourse (0/3, 0%). Five of the six participants reported receiving oral sex (**Supplemental Table 6**).

Significantly higher median JSD values of the MSU, but not peri-urethral, microbiotas were associated with participant-reported sexual activity for three of the six applicable participants: ProFUM05, ProFUM06, and ProFUM08 (**Supplemental Table 7A**). We next separated the responses by type of sexual activity; four of the six applicable participants (ProFUM02, ProFUM05, ProFUM07, and ProFUM08) had significant associations between MSU microbiota JSD values and vaginal intercourse, while only two participants (ProFUM02 and ProFUM08) had an association with oral sex (**Table 3**). However, the two participants with significant associations between MSU microbiota JSD values and oral sex were the only two participants who always co-reported oral sex with vaginal intercourse, suggesting that the trend with sexual activity may be due to vaginal intercourse. To determine the nature of MSU microbiota variability associated with vaginal intercourse, we assessed for differences in microbiota composition and diversity between days with and without vaginal intercourse reported. Median alpha-diversity measures following vaginal intercourse were significantly higher for two participants (**Supplemental Table 7B**). Four participants had at least one bacterial taxon that was statistically significant (**Supplemental Table 5B**). The exceptions were participants ProFUM02 and ProFUM06; however, since these participants only reported vaginal intercourse on two and one day(s), the power of analysis to detect significant changes in the taxa was low. Participants ProFUM04, ProFUM05, ProFUM07, and ProFUM08 all had significantly higher frequencies of detection of various *Streptococcus* and *Staphylococcus* species following vaginal intercourse. Participant ProFUM02 also had higher frequencies of *Streptococcus* species, but this was not significant (*p*=0.081). **Supplemental Table 5B** also shows that some species, none of which belong to the *Streptococcus* or *Staphylococcus* genera, had significantly lower frequencies of detection following vaginal intercourse. These data may affect the alpha-diversity values seen in **Supplemental Table 7B**. Overall, changes in MSU microbial composition were associated with vaginal intercourse, but unlike with menstruation, we observed similar trends both within and among participants.

**Table 3.** Association between Jensen-Shannon Divergence Values for MSU Microbiota and Participant-reported Vaginal Intercourse and Oral Sex.

| MSU Microbiota (Median JSD Values) |  |  |  |  |  |  |
| --- | --- | --- | --- | --- | --- | --- |
| Participants | Penetrative Vaginal Intercourse |  |  | Receiving Oral Sex |  |  |
|  | "Yes" (n) | "No" (n) | p-value | "Yes" (n) | "No" (n) | p-value |
| **ProFUM01** | N/A | N/A | - | N/A | N/A | - |
| **ProFUM02** | 0.559 (2) | 0.136 (64) | <0.001 | 0.559 (2) | 0.136 (64) | <0.001 |
| **ProFUM03** | N/A | N/A | - | N/A | N/A | - |
| **ProFUM04** | 0.057 (27) | 0.063 (45) | 0.373 | N/A | N/A | - |
| **ProFUM05** | 0.200 (21) | 0.108 (46) | 0.036 | 0.264 (4) | 0.125 (63) | 0.078 |
| **ProFUM06 (16S)** | 0.173 (1) | 0.123 (65) | 0.152 | 0.150 (2) | 0.123 (64) | 0.183 |
| **ProFUM07** | 0.242 (4) | 0.057 (65) | <0.001 | 0.163 (4) | 0.057 (65) | 0.094 |
| **ProFUM07 (16S)** | 0.057 (4) | 0.054 (65) | 0.150 | 0.083 (4) | 0.054 (65) | 0.117 |
| **ProFUM08** | 0.279 (3) | 0.071 (64) | <0.001 | 0.279 (3) | 0.071 (64) | <0.001 |

Median JSD values shown for MSU microbiota on days when the participant reported ("Yes") or did not report ("No") vaginal intercourse (*left*) or oral sex (*right*). Mann-Whitney U test used to determine significance. *p*-value < 0.05 is significant (green). N/A (Not applicable) refers to participants who did not report the personal factor.

### Assessing for Confounding Factors

We next determined if the described trends were confounded by other reported factors. Tampon and or pad use nearly always co-occurred with menstruation, as did daily number of bowel movements for three participants (**Supplemental Table 8A**), while oral sex frequently co-occurred with vaginal intercourse (**Supplemental Table 8B**).

### Urine Property Dynamics

To determine if the described trends resulted from an altered urinary environment, we assessed for relationships between personal factors and urine properties, as measured by a urine dipstick. With few exceptions, blood was the only significantly altered dipstick-measured urine property during menstruation (**Table 4A**). No changes in any of the dipstick measures were associated with vaginal intercourse (**Table 4B**).

**Table 4.**
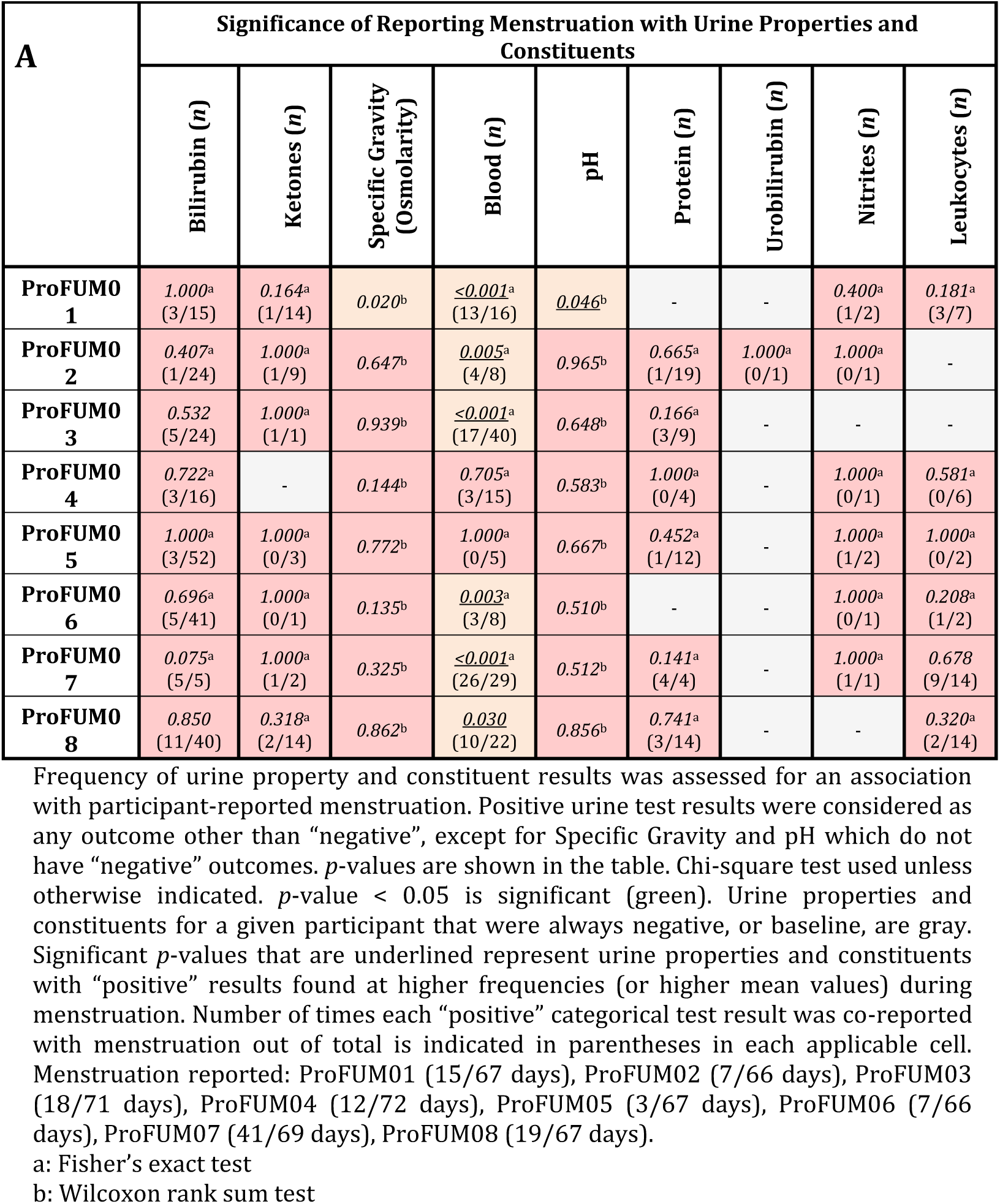
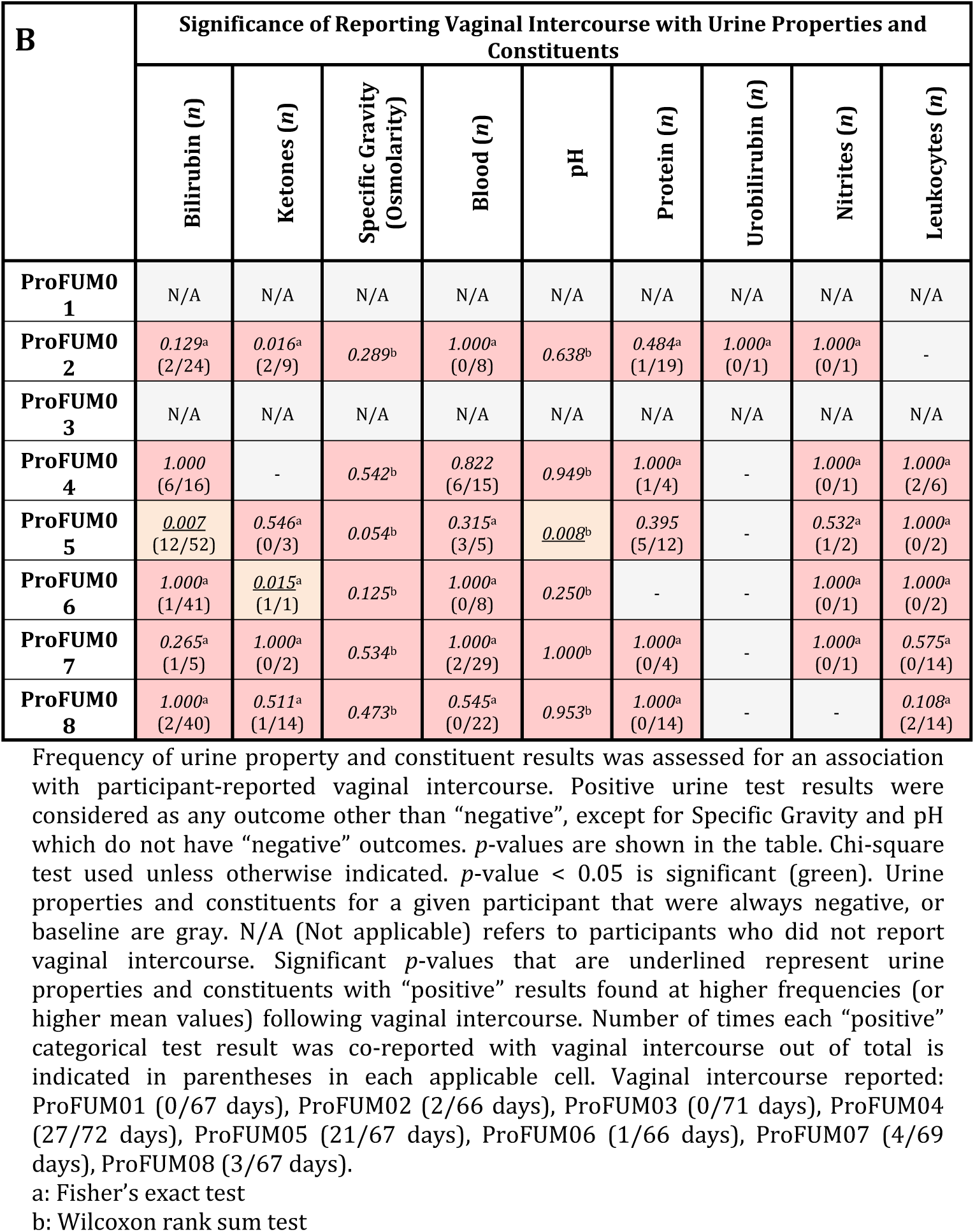
Significance of Participant-reported Personal Factors with Dipstick-Measured Urine Property Results.

## DISCUSSION

This study shows that the LUT microbiota is both dynamic and resilient. Our findings demonstrate variability in the voided urine sample of adult women participants, revealing the dynamic nature of the LUT microbiota and providing evidence that both menstruation and sexual activity influence those dynamics. They also show that the LUT and peri-urethra are distinct microbiological niches in most young women.

Using JSD, an analytic approach commonly used to measure longitudinal stability, we found that, within an individual, the peri-urethral microbiota is more variable than the LUT microbiota. Physiologically, this is not surprising, as the peri-urethral area is exposed to the genital and external environment, while the LUT is not. Thus, transitions of flora may occur more frequently in the peri-urethra than the LUT.

Because post-urethral (e.g. vulva and/or vaginal) microbes are often present in MSU samples^35 37^, it was essential to include a control for specimen quality, which is why we used a peri-urethral swab to measure specimen collection compliance. This peri-urethral control allowed us to make conclusions that exclusively relate to the LUT microbiota rather than to those of the peri-urethra. Since we did not collect vaginal swabs, we cannot comment on bacterial interplay between the peri-urethra and vagina. As this is the first study to assess peri-urethral microbiota temporal dynamics, we cannot compare our findings to others. It is possible that peri-urethral microbiota variability relates to specimen collection variability. Participants were instructed verbally and visually to collect the peri-urethral specimens by swabbing 1cm lateral to the urethral opening. However, one would expect that with repeated collection over three months, the participant’s technique to collect the peri-urethral specimen would become more standard and consistent, meaning JSD values would become lower (i.e., more stable) throughout the study. Qualitatively, this did not occur, suggesting that the daily variability of the peri-urethral microbiota is biologically meaningful and not likely due to collection inconsistencies.

Sexual activity was associated with increased variability in the MSU – and the type of sexual activity mattered. Moreover, this sex-associated variability differed within individuals. The elevated frequency and abundance of *Streptococcus* and *Staphylococcus* species in the MSU specimens were consistent findings following vaginal intercourse. One might predict microbiota variability to be related to oral sex. Indeed, participant ProFUM04, who only reported vaginal intercourse (27/72 days), did not have significantly different JSD values for either MSU or peri-urethral microbiotas (**Table 3**). Furthermore, a relationship between variability and oral intercourse might be supported by the fact that *Streptococcus* species are commonly part of the oral flora^38^. However, if one considers the other participants’ data, this relationship is less convincing. For example, participant ProFUM05 reported sexual activity 21/67 days; 17/21 days she reported vaginal intercourse only, while the other 4/21 days she reported both vaginal and oral sex. Assessment of these latter four days alone shows no significant association with MSU microbiota stability, while assessment of all 21 days does (**Table 3**). Furthermore, participant ProFUM07 reported sexual activity 6/69 days; 2/6 days she reported only vaginal intercourse, 2/6 only oral sex, and 2/6 both. Again, only an assessment of days including vaginal intercourse (4/6) showed significant associations with MSU microbiota stability (**Table 3**). Altogether, these data suggest that vaginal intercourse, rather than oral sex, is associated with MSU microbiota variability.

In the three participants who were asked (i.e., ProFUM06, ProFUM07, and ProFUM08), condom use was reported 100% of the time when vaginal intercourse was reported. Therefore, it is impossible to make a conclusion regarding the influence of condom use on the relationship between MSU microbiota variability and vaginal intercourse from these data alone. Sexual partner variability was different among the applicable participants; whereas participant ProFUM07 reported the same male sexual partner for each instance of vaginal intercourse (N=4), participant ProFUM08 reported different male partners (N=3). To conclusively show a role for the male sexual partner in the MSU microbiota variability of the female, it would be necessary to show movement of genetically related isolates between the partners following vaginal intercourse longitudinally. This has been demonstrated by Eren and co-workers, who observed strong correlations between unique sequence variants of *Gardnerella vaginalis* strains obtained from vaginal and urethral/penile samples of sexual partners^39^.

The effects of vaginal intercourse on the vaginal and MSU microbiotas appear to differ. Disruptions in the stability of the vaginal microbiota have been associated with vaginal intercourse^3^. This includes an increase in *Gardnerella*^3 40 41 42^ sometimes accompanied by a decrease in *Lactobacillus*, particularly *L. iners*^40^ and *L. crispatus*^42^. But this increase is not seen when condoms are used^40^. The leading theory of the mechanism behind these changes is that the altered microbiota comes from the male ejaculate, which contains microbes of the male urethra. The microbiome of semen has been very recently characterized and found to contain abundances of bacteria, many of which are part of the vaginal microbiota (e.g., *Lactobacillus, Veillonella, Streptococcus, Porphyromonas, Atopobium*), including *Gardnerella*^42^. Mandar et al. recently assessed 23 heterosexual couples and found that the semen microbiota, collected several days prior to intercourse, was more similar to the partner’s vaginal microbiota after vaginal intercourse^42^. We did not observe increases in *Gardnerella* in the MSU specimens following vaginal intercourse, but rather increases in *Streptococcus* and *Staphylococcus. Streptococcus* is a predominant taxon in the semen microbiota^42^. However, we observed these trends even when condom use was reported (e.g., ProFUM07 and ProFUM08) and therefore the bacteria are not likely from the male ejaculate. Overall, the only comparable finding is that both the vaginal and MSU microbiota have decreased stability following vaginal intercourse, but the mechanisms underlying these changes are likely very different, which is likely a reflection of the unique physiology of each site.

Menstruation (and associated activities, such as menstrual products) increased variability. Whereas our data revealed trends with obvious variability (i.e. JSD values), they were highly individualized. Thus, it is difficult to determine the mechanistic relationship between menstruation and MSU microbiota variability. The microbes found at higher frequencies and abundances during menstruation (e.g., *Corynebacterium, Staphylococcus, Actinomyces*) are primarily skin flora^1^. One possible explanation might be exogenous introduction via feminine hygiene product use, which was reported at significantly higher frequencies during menstruation for all participants (**Supplemental Table 10**). However, if hygiene products introduced bacteria to the LUT, one may expect similar changes to the peri-urethra, yet we observed no change in peri-urethral microbiota stability with menstruation or hygiene product use. An alternative explanation might be that the LUT environment changes during menstruation, thus favoring outgrowth of different microbes. Except for blood, however, there were no significant changes in the frequency of positive urine property and constituent results during menstruation (**Table 4**). The blood likely originates from the vaginal tract and thus is not directly relevant to the LUT environment; yet, this serves as an important positive control to verify the integrity of participants’ responses to the questionnaire. In addition to feminine hygiene product use, only reported number of bowel movements was found to have a significant association with menstruation (**Supplemental Table 8A**). However, when we assessed for associations between number of bowel movements and microbiota characteristics, we found no significant trends. Hormonal effects are also a possibility but were not investigated here.

While we relied primarily on EQUC, we showed that those results are comparable to those obtained with 16S rRNA gene sequencing. We analyzed MSU specimens from ProFUM07 by both modified EQUC and 16S rRNA gene sequencing. (**Figures 3G & 3H**, respectively). Qualitatively, the microbiota and microbiome were dominated by *Lactobacillus* (blue) except for three, one to two-week periods during the beginning (i.e., week 1), middle (i.e., weeks 3 and 4), and end (i.e., weeks 7 and 8). This deviation from *Lactobacillus* dominance occurred for the same MSU specimens by both methods. Appropriately, JSD values were elevated for both methods during these periods. In fact, JSD values for the two methods followed an almost identical pattern over time (**Supplemental Figure 2**). These data show that the two methods are complementary and validate the use of only one method to analyze the specimens of the other seven participants.

We have limited our approach to assessing the relationship between personal factors and the microbiota of the subsequent day. It is difficult to discern whether it would be biologically appropriate to further expand the analyses. It is possible that some personal factors take longer than one day to significantly impact the microbiota. In previous studies, correlations between personal factors and UTI risk for example were studied across weeks, months, and even years, and thus do not provide relevant information regarding the timeliness of their effects. Nonetheless, our dataset represents a novel means to determine if relationships between personal factors and delayed changes to the LUT microbiota exist and is an important future direction to consider.

This study shows that it is feasible to perform longitudinal urine specimen collection in pre-menopausal sexually active women, although our “screen” for sample concordance excluded 1/3 of potential participants. This screening threshold may require revision with subsequent studies, as it may have biased our participant population; excluding participants based on lack of distinct microbiota between paired specimens may not actually relate to specimen collection compliance. Although others have suggested that the microbes of the peri-urethra and MSU are distinct^43 44 45^, rigorous and properly controlled studies assessing this distinction have yet to be performed. Our data show that these two niches are distinct in some women.

## METHODS

### ProFUM Study Enrollment and Specimen Collection

From September 2017 to July 2018, we invited asymptomatic pre-menopausal female employees from the Loyola University Medical Center campus to participate in an IRB-approved clinical trial (NCT03250208) entitled “Probiotics and the Female Urinary Microbiome (ProFUM) study.” The study was divided into three phases. Phases I and III flanked an experimental phase (i.e., Phase II), in which the participants were randomized to take an oral probiotic or placebo. This part of the study has been published^34^. MSU, peri-urethral swabs, and a personal questionnaire were collected daily throughout all three phases.

Using IRB-approved invitation methods, potentially interested individuals contacted a research nurse and were screened for eligibility during an in-person visit. Inclusion criteria were female gender at birth, over 18 years of age, and ability to read English and sign a consent form detailing the requirements and voluntary nature of the study. Exclusion criteria were current pregnancy, antibiotic or probiotic usage, or the plan to vacation for more than seven days during the time of specimen collection (i.e., three months). Individuals who met these eligibility criteria signed a consent form and were instructed that a 3-day sample collection screening was required prior to final enrollment in the study. This is described below.

Participants were given sufficient supplies (described below) and instructed on how to collect, label and deliver daily MSU and peri-urethral swab specimens to the research team. Participants were instructed on proper specimen collection through use of a standardized video detailing proper collection of an MSU specimen and a peri-urethral swab. Specifically, participants were instructed not to use the genital cleansing wipe provided in the urine collection kits. Participants were assigned unique study identifiers by the research nurse and were instructed to attach labels with their study ID and date of collection to each specimen prior to delivery to a locked drop-box in an accessible room at Loyola University Medical Center.

MSU specimens were collected by voiding into a toilet to discard the initial void (i.e., approximately the first 10 mL of urine). The remaining specimen was then collected into a sterile collection cup. A portion of each urine sample was placed in a sterile manner into a BD Vacutainer® Plus C&S Preservative Tube for culturing. The tube contained a lyophilized boric acid preservative to prevent bacterial overgrowth without causing cell death, allowing specimens to be held at room temperature for up to 72 hours without altering integrity. Specimens that were collected more than 72 hours prior to receipt were excluded from analyses.

Peri-urethral swab specimens were collected using a BD ESwab® Liquid Amies Collection and Transport System. These flocked swabs allow for optimal elution of the specimen into the medium. Specimens were stored at room temperature. Due to lack of a preservative, specimens that were collected more than 48 hours prior to receipt were excluded from analyses due to the possibility of bacterial overgrowth and subsequently inaccurate data. Peri-urethral swabs were collected by swabbing vertically top to bottom approximately 1 cm lateral to the urethral opening.

Study specimen collection was initiated with a three-day assessment to verify proper specimen collection. Participants were withdrawn from further study participation if their MSU and peri-urethral specimens had similar microbial contents, as determined using the Bray-Curtis Dissimilarity test. A value of 0.8 was the threshold for eligibility. Participants continued in the study when their paired specimens with scores >0.8 for two of three collection days of the screening period. Continuing participants were scheduled for a second one-on-one meeting with the clinical team during which the continuing research tasks associated with the ProFUM study were described and participants completed a demographics questionnaire, which included: age, race/ethnicity, height/weight, blood pressure, vaginal parity, birth control method, condom use, typical length of menstrual cycles, use of menstrual hygiene products, prior urogynecologic surgery, sexual activity (frequency, type, partners), dietary preferences, alcohol consumption, number of bowel movements in an average week, use of cigarettes, frequency of bathing, current medications, and history of UTI, kidney stones, urinary incontinence, and fecal incontinence.

In addition to specimen collection, participants completed a daily personal questionnaire regarding life events and behaviors of the previous 24 hours. The non-validated questionnaire queried alcohol consumption, medications, medical events, menstruation, menstrual hygiene, bathing, swimming, sexual activity (oral, vaginal, other), and number of bowel movements. The questionnaire included two yes/no-style questions regarding urination and bowel movements for the current day (i.e., since waking up and before specimen collection). The last three participants completed a modified questionnaire that included two additional questions regarding condom use with vaginal intercourse and whether the sexual partner had changed. Completed questionnaires were given to the research team with each specimen.

### Laboratory Analysis

A modified version of EQUC protocol was conducted to identify microbiota^27^. For the MSU specimens, 0.01 mL of urine was spread quantitatively onto diverse media types [BAP, CNA agar, and CDC anaerobe 5% BAP (ABAP)] and incubated in appropriate environments at appropriate temperatures (5% CO2 at 35°C for 48 hours or anaerobic conditions at 35°C for 48 hours). Each morphologically distinct colony type was counted and isolated to prepare a pure culture for identification by MALDI-TOF MS, using the Bruker MALDI Biotyper Research (RUO) System. All swab specimens underwent the same protocol using 0.01 mL of the liquid elution media after the swab was vortexed in the collection tube for 10 seconds.

Each MSU specimen was characterized using a urine dipstick. Approximately 1 mL of urine was pipetted onto a Siemens Multistix® 10 SG Reagent Strip. The reagent strip tests for the presence and quantity (or relative quantity) of glucose, bilirubin, ketones, hemoglobin (i.e., blood), protein, urobilinogen, nitrites, and leukocytes. These strip tests also measure pH and specific gravity. Results were read and interpreted according to manufacturer’s instructions.

The remaining volumes of the MSU and swab specimen elution media after bacterial culture were aliquoted for 16S rRNA gene sequencing. The nucleic acid preservative AssayAssure (Sierra Molecular, Incline Village, NV) was added (10% relative to specimen volume) before storage at -80°C.

DNA isolation, polymerase chain reaction (PCR) amplification, and 16S rRNA gene sequencing of urine cultures has been described^46^. To minimize contamination, isolation of DNA was performed in a laminar flow hood. Genomic DNA was extracted from MSU and swab elution media with the Qiagen DNeasy Blood and Tissue kit. 1 mL of urine or 0.5 mL of swab elution media was used. Peptidoglycan degrading enzymes, mutanolysin and lysozyme, were added to ensure robust lysis of Gram-positive and Gram-negative species^47^. DNA was eluted into 50 µl of buffer AE, pH 8.0, and stored at -20° C. The hyper-variable region 4 (V4) of the bacterial 16S rRNA gene was amplified via a two-step PCR protocol^27 31^. The PCR reaction was purified and size-selected using Agencourt AMPure XP-PCR magnetic beads (Beckman Coulter, Pasadena, CA). Each sample was quantified using the Qubit fluorometeric system (Thermo-Fisher, Waltham, MA). The samples were pooled, quantified to a standard volume, and placed in the 2X 250 bp sequencing reagent cartridge, according to manufacturer’s instructions (Illumina, San Diego, CA).

After sequencing, sample barcodes and sequencing primers were removed using Illumina proprietary MiSeq post-sequencing software. The mothur program (v1.41.3) was used to process raw sequences following the recommended MiSeq standard operating procedure^48^. Briefly, mothur produced 16S contigs by combining paired end reads based on overlapping nucleotides in sequence reads; contigs of incorrect length for the V4 region (<290 bp, >300 bp) and/or contigs containing ambiguous bases were removed. Chimeric sequences were removed using UCHIME within the mothur package^49^. Subsampling at a depth of 5000 sequences was performed to correct for the different sequencing depths of each sample. The sequences were clustered into species-level operational taxonomic units (OTUs) with identity cutoff at 97%^50^. The OTUs were classified using RDP classifier (v2.11) at the genus level^50^ and BLCA^51^ at the species level.

### Statistical Analyses

No *a priori* power calculation was performed to determine appropriate sample size for this exploratory study; sample size was determined by feasibility, research team availability and budgetary constraints.

Statistical analyses of microbiota data were performed using SAS software version 9.4 (Cary, NC). Microbiota stability measures were calculated using Jensen-Shannon Divergence (JSD) as described^2^. First, we calculated a representative microbiota distribution for each participant, averaging the abundance data for the microbiota across all collection days. JSD values were then calculated between each day’s microbiota and the average microbiota for each participant. These values were then applied to metadata, such as questionnaire results, to determine if a statistical relationship exists between microbiota stability and personal factors. Wilcoxon rank sum tests and Mann-Whitney U tests compared mean or median JSD values with participant-reported personal factors. Frequency of bacterial detection was compared to personal factors using either Pearson chi-square or Fisher’s exact tests, depending on assumption validity. Chi-square testing was used to compare categorical variables. One-way analysis of variance (ANOVA) was used to compare continuous variables. Correlations between variables were determined using the Pearson Correlation test. All test results were considered significant using a p-value of <0.05. Relative abundance graphs, alpha diversity measures, and PCoA plots were generated in RStudio.

## Data Availability

Data is available upon request

## ACKNOWLEDGMENTS

We thank Mary Tulke RN for her assistance with participant recruitment. We also acknowledge funding from NIH (R01 DK104718 awarded to AJW and LB). The funders did not play a part in the design or conduct of the study.

TKP, BW, LB, ERM, and AJW designed the study. TKP and BW recruited participants. TKP processed the specimens. TH prepared them for DNA sequencing. RL sequenced the specimens. TKP, BW, QD and AJW analyzed the data. TKP and AJW wrote the manuscript. TKP, BW, TH, RL, LB, QD, ERM, and AJW reviewed and edited the manuscript.

## SUPPLEMENTAL FIGURE LEGENDS

**Supplemental Figure 1. Distribution of Jensen-Shannon Divergence Values for MSU and Peri-urethral Microbiota from all ProFUM Participants**. Box plots depict the range of JSD values of microbiota (EQUC of ProFUM 1-5, 7-8) and microbiome (16S rRNA gene sequencing of ProFUM06 and ProFUM07) data of MSU (**A**) and peri-urethral (**B**) specimens from all applicable ProFUM participants. Median scores are depicted by the solid lines in each box. The distribution of raw values are depicted by the overlaid circles.

**Supplemental Figure 2. JSD Values for MSU Microbiota of Participant ProFUM07 Obtained using Culture and Sequencing**. JSD values were calculated for MSU microbiota of participant ProFUM07. Microbiota data was obtained using modified EQUC (solid line) and 16S rRNA gene sequencing (dashed line). Data are plotted over time in days.

## SUPPLEMENTAL TABLES

**Supplemental Table 1. Bray-Curtis Dissimilarity Values for Paired Specimens during the Screening Phase**.

**Supplemental Table 2. Overview of Bray-Curtis Dissimilarity Values Comparing MSU and Peri-urethral Microbiota of the ProFUM Participants based on EQUC data**.

**Supplemental Table 3. Qualitative Description of the Microbial Temporal Dynamics of MSU Specimens of the ProFUM Participants shown in Figure 3**.

**Supplemental Table 4. Association between Alpha-Diversity Values for MSU Microbiota and Participant-reported Menstruation**.

**Supplemental Table 5. List of MSU Taxa with Significantly different Frequencies of Detection during (A) Participant-reported Menstruation; (B) Participant-reported Vaginal Intercourse**.

**Supplemental Table 6. Summary of Participant-reported Personal Factors Relating to Sexual Activity**.

**Supplemental Table 7 (A) Association between JSD Values for MSU and Peri-urethral Microbiota and Participant-reported Sexual Activity. (B) Association between Alpha-Diversity Values for MSU Microbiota and Participant-reported Vaginal Intercourse**.

**Supplemental Table 8 (A) Significance of Reporting Menstruation with Potentially Confounding Personal Factors. (B) Significance of Reporting Vaginal Intercourse with Potentially Confounding Personal Factors**.

